# Measuring the influence of expectations, beliefs, and medication side effects on the risk for drug discontinuation among individuals starting new medications

**DOI:** 10.1101/2023.09.18.23295758

**Authors:** David F Blackburn, Shenzhen Yao, Jeffery G. Taylor, Qais Alefan, Lisa M. Lix, Dean T. Eurich, Niteesh K. Choudhry

**Author notes:** Correspondence to: David Blackburn, College of Pharmacy and Nutrition, University of Saskatchewan, 2A20.20 Health Sciences, 107 Wiggins Road, Saskatoon, SK, S7N 5E5. Contributor and guarantor information: The corresponding author attests that all listed authors meet authorship criteria and that no others meeting the criteria have been omitted David Blackburn: Conceptualization, Methodology, Analysis, Writing – Original draft preparation, Project administration, Fund acquisition. Qais Alefan: Analysis, Project administration, Writing. Shenzhen Yao: Methodology, Analysis, Writing. Jeffrey G. Taylor: Methodology, Writing – Original draft preparation. Lisa M. Lix: Methodology, Analysis, Writing. Dean T. Eurich: Methodology, Analysis, Writing. Niteesh K. Choudhry: Conceptualization, Methodology, Analysis, Writing. Competing interests: *Blackburn*, *Taylor*, *Eurich*, *Lix*, and Choudhry received grant funding from the Canadian Institutes of Health Research (CIHR grant number 130343) for the submitted work; no financial relationships with any organizations that might have an interest in the submitted work in the previous three years; no other relationships or activities that could appear to have influenced the submitted work. Disclaimer: This Study is based in part on de-identified data provided by the Saskatchewan Ministry of Health. The interpretation and conclusions contained herein do not necessarily represent those of the Government of Saskatchewan or the Saskatchewan Ministry of Health.

## Abstract

**Objectives:** To measure the impact of beliefs, expectations, side effects, and their combined effects on the risk for medication non-persistence.

**Design:** A population-based questionnaire.

**Setting and Participants:** Individuals from Saskatchewan, Canada who started a new antihypertensive, cholesterol-lowering, or antihyperglycemic medication were surveyed about risk factors for non-persistence including: (a) beliefs measured by a composite score of three questions asking about the threat of the condition, importance of the drug, and harm of the drug; (b) incident side effects attributed to treatment; and (c) expectations for side effects before starting treatment. Descriptive statistics and logistic regression models were used to quantify the influence of these risk factors on the outcome of non-persistence. Odds ratios (OR) and 95% confidence intervals (CI) were estimated.

**Main Outcome Measure:** Self-reported medication non-persistence.

**Results:** Among 3,029 respondents, 5.9% (n=179) reported non-persistence within four months after starting the new drug. After adjustment for numerous covariates representing socio-demographics, healthcare providers, medication experiences and beliefs, both negative beliefs (OR 7.26, 95% CI: 4.98 to 10.59) and incident side effects (OR 8.00, 95% CI 5.49 to 11.68) were associated with the highest odds of non-persistence with no evidence of interaction. In contrast, expectations for side effects before starting treatment exhibited an important interaction with incident side effects following treatment initiation. Among respondents with incident side effects (n=741, 24.5%), the risk for early non-persistence was 11.5% if they indicated an expectation for side effects before starting the medication compared to 23.6% if they did not (adjusted OR 0.38, 95% CI 0.25 to 0.60).

**Conclusions:** Expectations for side effects may be a previously unrecognized but important marker of the probability to persist with treatment. A high percentage of new medication users appeared unprepared for the possibility of side effects from their new medication making them less resilient if side effects occur.

**What is already known on this topic:**

- Prior expectations for side effects are thought to increase the risk for nocebo effects and increase the risk for medication non-persistence.
- Medication non-persistence remains a major threat to patient outcomes.

**What this study adds:**

- Expectations for side effects from medications may be a previously unrecognized *protective* factor against non-persistence.
- A high percentage of new medication users appear unprepared for the possibility of side effects from their new medication making them less resilient if side effects occur.

---

Cardiovascular diseases consume a major portion of total health care expenditures (i.e., $350 billion annually in the USA).^1^ Although increased use of risk-modifying medications has contributed to declining morbidity and health-spending over the past two decades, adherence to these medications remains low.^2-5^ Multiple studies have documented the high prevalence of non-adherence to cardiometabolic medications^2-5^ resulting in poor risk factor control,^6,7^ hospitalizations,^8,9^ and potentially avoidable deaths.^10,11^

Non-persistence to medications often occurs within days to weeks after the first dose.^4,12,13^ Although side effects are considered a major cause of early non-persistence,^14^ the willingness to persist with a new medication is highly influenced by an individual’s assessment of necessity versus concern.^15^ A minor side effect may be unbearable for a person who believes their drug has few health benefits, while severe side effects may be tolerated by those believing their drug is essential. We previously found that negative medication beliefs were the most powerful predictors of early drug discontinuation among people experiencing side effects.^16^ That is to say, patients with low confidence in their medication were more likely to discontinue it after a side effect occurred. Notably, expectations and beliefs can also influence the perception of side effects in addition to influencing the response to them.^17-21^ However, it is unknown if heightened perceptions of side effects (i.e., the nocebo effect) will increase non-persistence to a greater extent than what would be expected from negative beliefs alone. The practical relevance of these unresolved issues faces health care providers daily. It could be argued that a prominent objective of many health care providers is to inform individuals about side effects that should be expected.^22^ Our research aim was to measure the impact of negative beliefs, expectations, incident side effects, and their combined effects on premature drug discontinuation (i.e., non-persistence).

## Methods

Study data were from a population-based questionnaire mailed to residents in the province of Saskatchewan, Canada. Saskatchewan has a universal drug insurance program that covers approximately 90% of the population of over one million residents regardless of age or socioeconomic status. All beneficiaries of the drug insurance program were eligible to receive the questionnaire if they were at least 30 years old upon receiving a new claim for an antihypertensive, cholesterol-lowering, or antihyperglycemic medication between September 2019 and February 2020. To restrict the sample to new users only, eligible beneficiaries required at least two years of continuous registration preceding the earliest claim without any prior claims for these eligible medications.^16^ Screening for eligible beneficiaries was conducted on several occasions during the two year period; thus, questionnaires were mailed within four to six weeks of the earliest dispensation for one of these eligible medications. The mailout and consent form followed the Dillman strategy including an invitation letter, questionnaire package, and follow-up reminder.^23^

### Participants

Respondents were included in the analysis if they: a) confirmed receiving one of the eligible medications within the past four months; b) returned a completed survey with a completed consent to participate; and c) answered all items relating to persistence (one question), side effects (one question), and medication beliefs (three questions). Questionnaire design and dissemination have been described in detail elsewhere.^16^ Briefly, the questionnaire contained 58 items designed to collect information on factors influencing adherence during the early phase of treatment including: patient-related factors (e.g., knowledge, attitudes, beliefs), social and economic factors (e.g., income, marital status), treatment-related factors (e.g., side effects), and health care system factors (e.g., appointment length, relationship/trust/support).^16^

Seven of the questionnaire items were grouped into two composite scores developed previously: a knowledge score and health care provider support score.^16^ The first score represented health care provider support and included five questions for a maximum of 25 points. Questions asked if respondents were given a chance to ask the doctor questions, received information about side effects, if the doctor spent time to help them understand, if a nurse or pharmacist spent time, and if they trusted the doctor. Higher scores represented greater support from a health care provider (Cronbach’s alpha 0.82).^16^ Also, a knowledge score was calculated from two questions asking if they knew what the medication was used for and the reasons why the medicine was good for them (Cronbach’s alpha 0.77).^16^ Both scores were dichotomized into binary variables as described previously.^16^

### Non-persistence, incident side-effects, expectations for side effects, and beliefs

The primary outcome was non-persistence, measured with the following question, “*Are you still taking the new medicine prescribed to you?”* (yes or no). Incident side-effects were identified by asking, “*Did you experience side effect(s) from your new medicine?”* (yes, no, not sure). Expectations for side effects was elicited by asking, “*You expected to get side effects from this new medicine before you started taking it*” (strongly agree, agree, not sure, disagree, strongly disagree).

Beliefs about medications were measured using three items with 5-point Likert scale response options ranging from strongly agree to strongly disagree. The first question asked about the ‘*perceived threat of the medical condition’* (i.e., “*Your new medicine is for a condition that is a danger to your health*”). The second question focused on the ‘*expected importance of the drug’* (i.e., “*You are convinced that your new medicine is important for your health*”), and the third asked about ‘*perceived drug harms’ (i.e.,* “*You worry that your new medicine will do more harm than good*”). These questions were based on previously published questionnaires^15,24^ and demonstrated concordance validity through a strong association with the risk of non-persistence among a subgroup of respondents who experienced side effects in our previous study.^16^ We calculated an overall beliefs score for each respondent based on the sum of responses to the three questions.^24^ The beliefs score produced a total possible score of 15 points and a minimum of three points. Based on the overall beliefs score, individuals were dichotomized into “positive beliefs” and “negative beliefs” using the 25^th^ percentile threshold (i.e., score of <11 was considered “negative beliefs”). Cronbach’s alpha^25^ for the overall beliefs score was 0.68.

### Statistical Analysis

We presented baseline characteristics descriptively for the overall sample and for persistent and non-persistent respondents using frequencies, percentages, means, standard deviations, and medians. Differences in baseline characteristics between those reporting persistence and non-persistence were tested using chi-squared statistics.

Multivariable logistic regression models were used to assess the impact of beliefs, manifest side effects, and expectations for side-effects on the outcome of non-persistence in addition to the entire array of possible adherence predictors in the questionnaire. We fit a series of models in which all variables, other than age and sex, were first tested independently with the outcome and included in the final model only if they reached a significance level p<0.10 on univariate analysis and improved model discrimination performance determined by a statistically significant improvement in the integrated discrimination improvement statistic (IDI).^26^ The final model also included age greater than 65 years and sex. None of the variables in the final model exhibited multicollinearity defined as a variance inflation factor of 2.5 or greater. We also tested a two-way interaction between side effects and beliefs to identify any statistical evidence of influence between these variables. We then tested the interaction between an “expectation for side effects” and manifest side effects. Finally, we repeated the logistic regression analysis using individual beliefs questions rather than the overall beliefs score to ensure our results were not influenced by the decision to represent beliefs with a single score.

A c-statistic (i.e., area under the receiver operating characteristic curve) was calculated for each model to assess overall discriminative performance.^27^ Odds ratios (ORs) and 95% confidence intervals (95% CIs) were estimated. All analyses were conducted using SAS Software v9.4 (SAS Institute Inc, Cary, NC, USA). Our study was approved by the University of Saskatchewan Committee for Ethics in Human Research.

### Patient and Public Involvement

During the development and pilot testing of our questionnaire, we sought feedback from patients on readability and acceptability of the tool. For the current manuscript, the study team had no representation from patient or public groups. However, the focus of this research was to collect and analyze information from patients on their experiences and attitudes regarding a new medication.

## Results

Out of 11,970 eligible individuals who received the survey, 3,973 responded yielding a response rate of 33.2%. Of these, 3,029 (76.2%) reported starting new antihypertensive, cholesterol-lowering, or antihyperglycemic medication and answered all questions relating to side effects and beliefs [Figure 1]. Respondents were equally distributed between males (50.1%) and females (49.9%) and the average age was 62.2 years (SD 11.7 years). The majority of respondents were Caucasian (90.4%, n=2,724), two-thirds had pursued formal education or training beyond high school (64.1%, n=1,934), and over three-quarters were married or living with a partner (77.3%, n=2,319). Most respondents listed general health status and mental health status as good to excellent (85.3% and 91.3% respectively), and 86.9% (n=2,510) earned more than $25,000 annually (CAD). Lipid-lowering medications were the most common new drug reported (53.5%, n=1,620), followed by antihypertensives (28.9%, n=876), and antihyperglycemics (17.6%, n=533). Almost three quarters of patients received the prescription from their ‘regular doctor’ (72.0%, n=2,180) and 78.6% (n=2,415) were taking two or more medications daily at the time of completing the questionnaire [Table 1].

**Figure 1.**
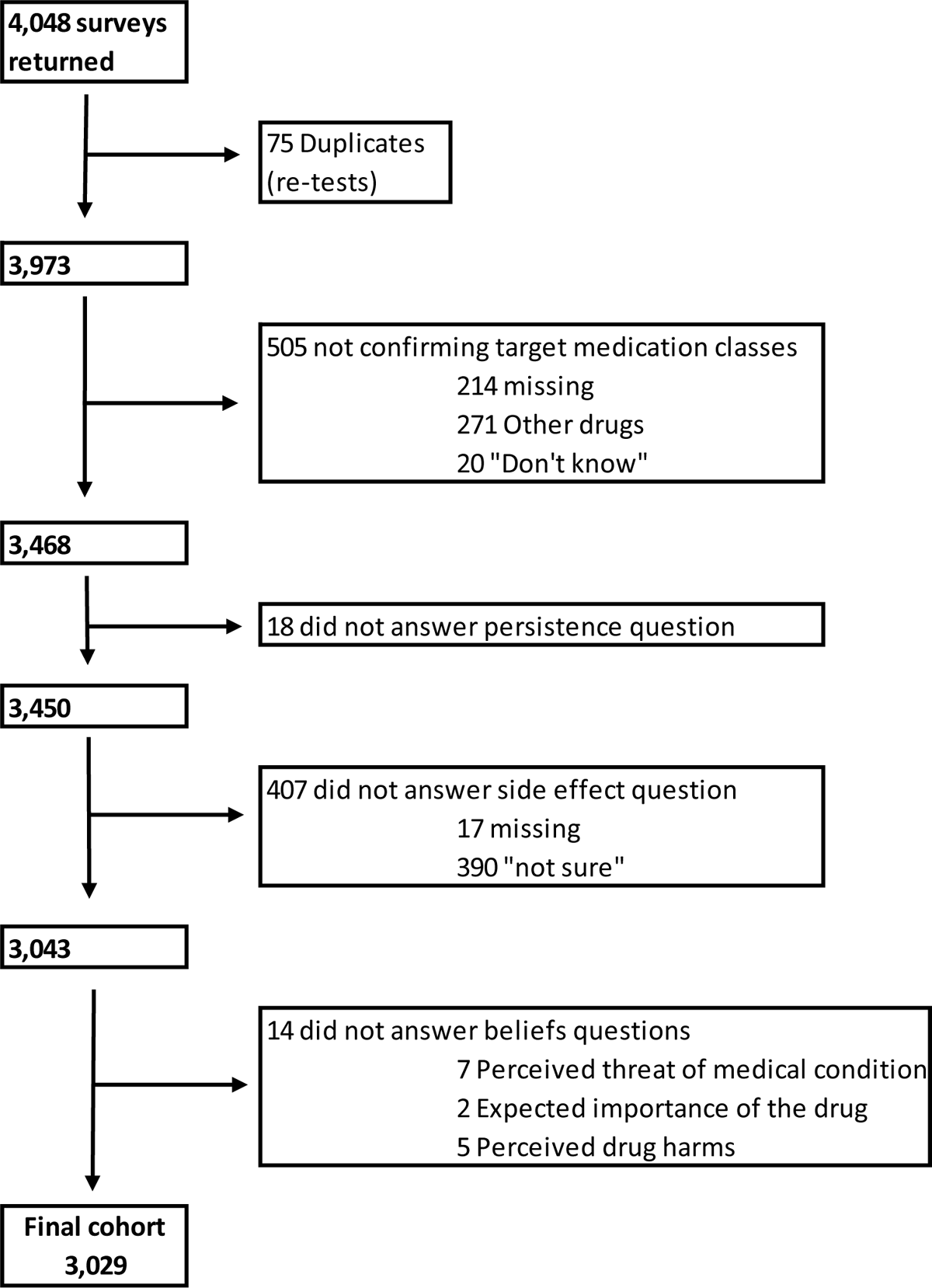
Flowchart for selection of study cohort

**Table 1.**
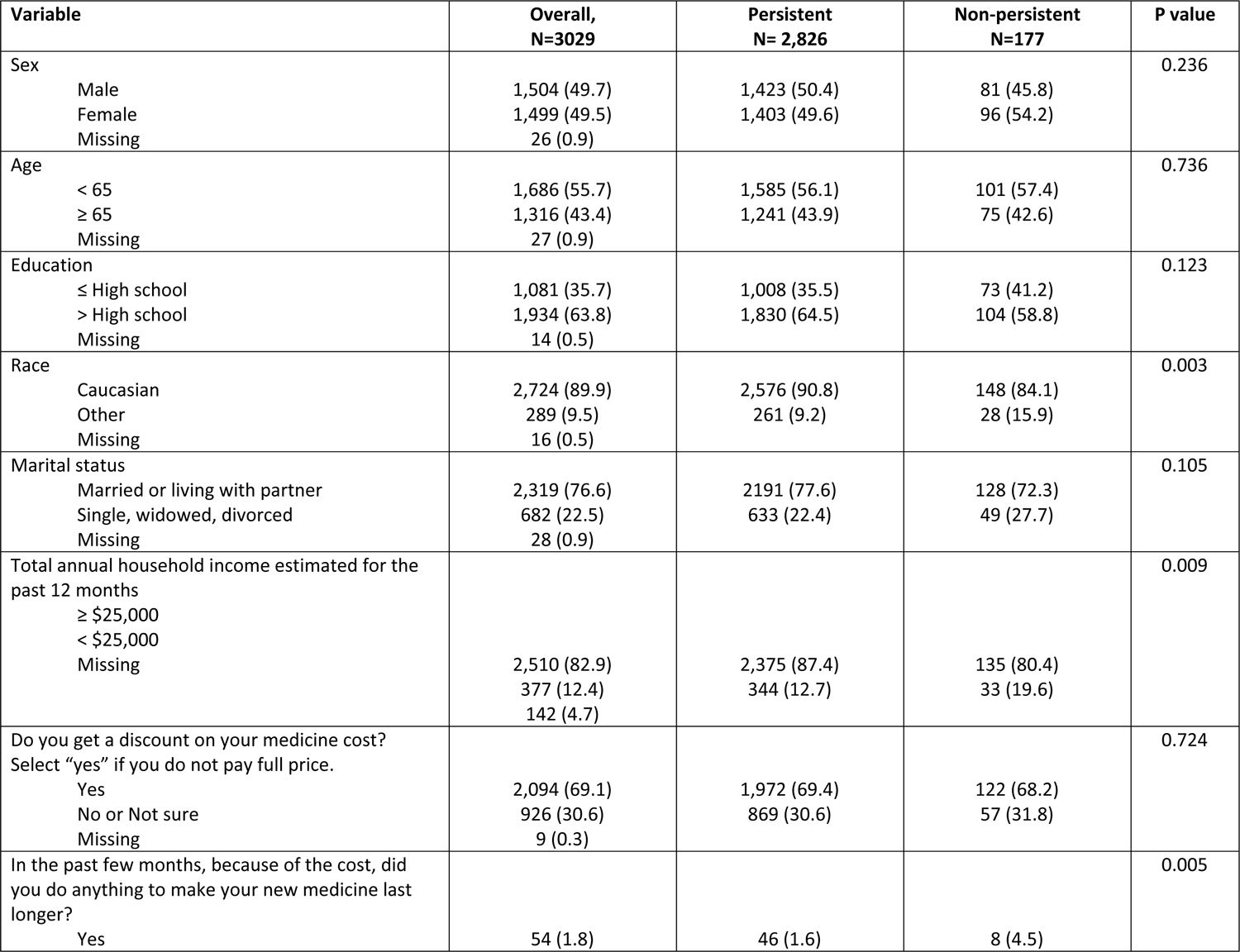
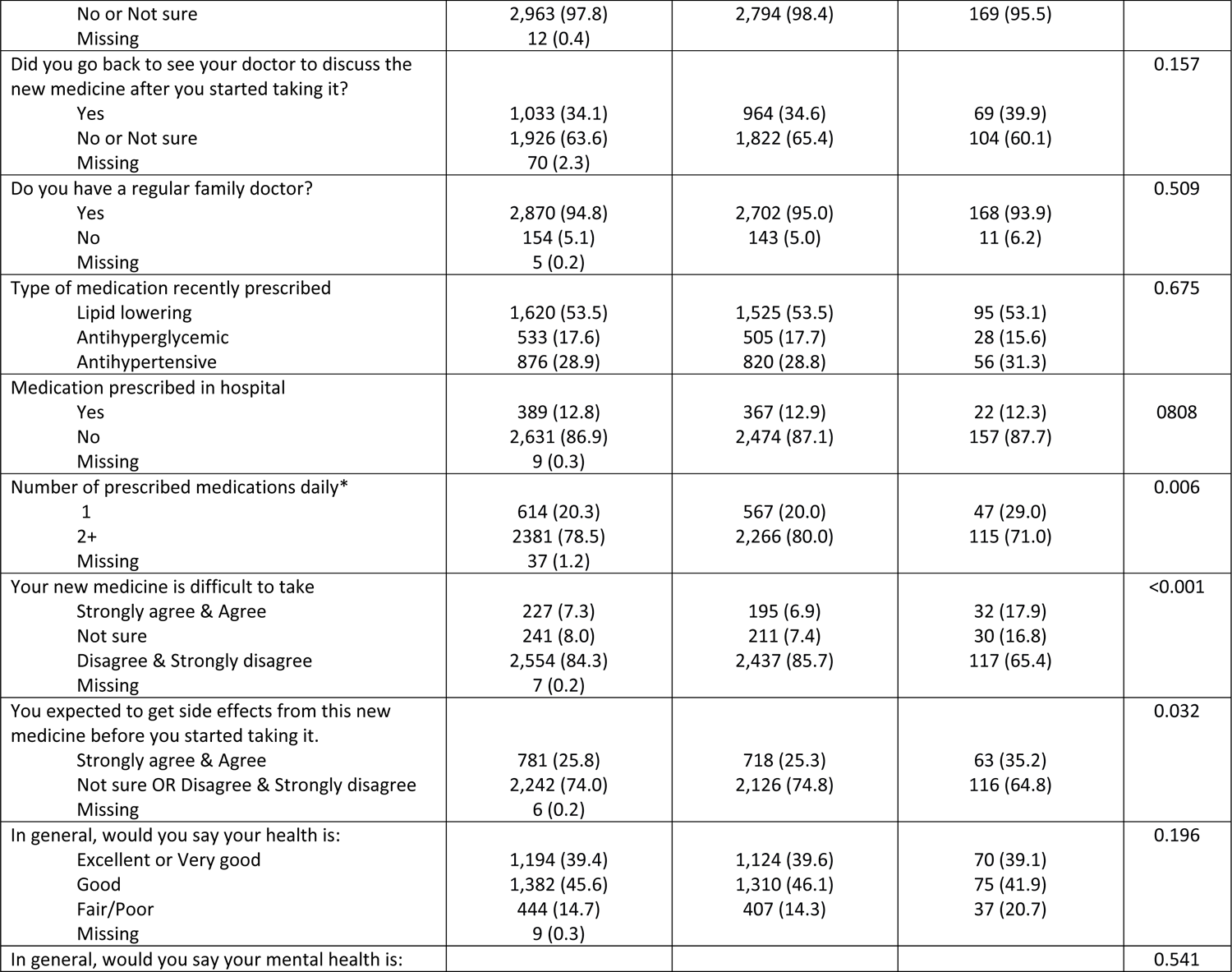
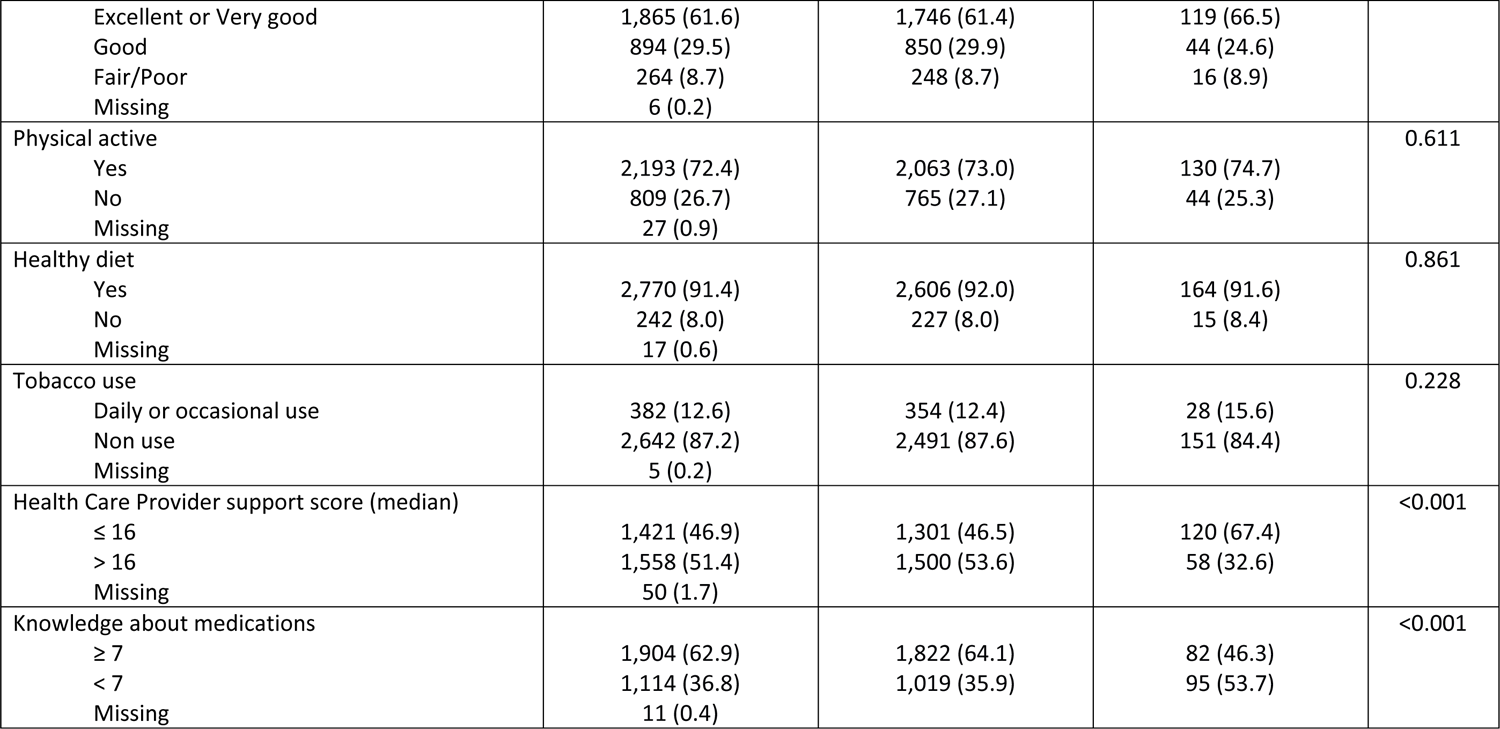
Baseline characteristics of the study cohort.

Negative beliefs were expressed by only 1.8% to 10.2% of respondents in each of the three beliefs questions. The median overall beliefs score was 12 (range 3 to 15) and respondents falling in the lowest quartile corresponded to a score <11 (17.2%, n=522). Incident side effects were reported by 24.5% (n= 741). One-quarter of respondents (25.8%, n=781) indicated they expected to get side effects from the new medicine before they started taking it.

Five variables were significantly associated with the outcome of non-persistence based on the univariate model results: negative versus positive beliefs (21.5% (n=112) vs 2.7% (n=67), p<0.001) [Table 2]; incident side effects versus no side effects (16.9% (n=125) vs 2.4% (n=54), p<0.001); expecting side effects before starting treatment (vs not expecting), (8.1% (n=63) vs 5.2% (n=116), p=0.032), a low (vs high) knowledge score (less than seven vs higher than six, 8.5% (n=95) vs 4.3% (n=82), p<0.001); and a low health care provider score (less than 17 vs higher than 16, 8.4% (n=120) vs 3.7% (n=58), p<0.001). The influence of side effects and beliefs on the risk for non-persistence were consistent regardless of the presence of each other.[Table 2] The highest risk of non-persistence was observed in people expressing both negative beliefs and side effects at the same time (38.9%, n=82), while the risk for non-persistence was extremely low in people without either risk factor (1.2%, n=24).[Table 2]

**Table 2.** Frequency and percentage of respondents expressing beliefs, side effects, and non-persistence.

| Item(s) | Response(s) | Percent of total group (n=3,029)(N) | % exhibiting non-persistence (i.e., row percentage)(n) | % of all non-persistence cases reported in total group (i.e., column percentage with denominator = 179)(N) | % experiencing side effects (i.e., row percentage)(N) | % exhibiting non-persistence among those WITH side effects (N) | % exhibiting non-persistence among those WITHOUT side effects(N) |
| --- | --- | --- | --- | --- | --- | --- | --- |
| Worry that med will do more harm than good | Negative belief (i.e., agree or strongly agree)* | 10.2 (309) | 30.7 (95) | 53.1 (95) | 55.3 (171) | 43.9 (75) | 14.5 (20) |
|  | Positive belief (i.e., disagree, strongly disagree, or unsure)* | 89.8 (2,720) | 3.1 (84) | 46.9 (84) | 21.0 (570) | 8.8 (50) | 1.6 (34) |
| Convinced med is important for your health | Negative belief (i.e., disagree, strongly disagree, or unsure)* | 1.8 (55) | 61.8 (34) | 19.0 (34) | 63.6 (35) | 74.3 (26) | 40.0 (8) |
|  | Positive belief (i.e., agree or strongly agree)* | 98.2 (2,974) | 4.9 (145) | 81.0 (145) | 23.7 (706) | 14.0 (99) | 2.0 (46) |
| Med is for danger condition | Negative belief (i.e., disagree, strongly disagree, or unsure)* | 4.9 (148) | 16.2 (24) | 13.4 (24) | 31.1 (46) | 37.1 (17) | 6.9 (7) |
|  | Positive belief (i.e., agree or strongly agree)* | 95.1 (2,881) | 5.4 (155) | 86.6 (155) | 24.1 (695) | 15.5 (108) | 2.2 (47) |
| Overall beliefs score derived from 3 questions above | Negative beliefs (<11) | 17.2 (522) | 21.5 (112) | 62.6 (112) | 40.4 (211) | 38.9 (82) | 9.7 (30) |
|  | Positive beliefs | 82.8 (2,507) | 2.7 (67) | 37.4 (67) | 21.1 (530) | 8.1 (43) | 1.2 (24) |

Side effects were reported more frequently in respondents with negative beliefs (i.e., 40.4% when beliefs score < 11 versus 21.1% when score was 11 or higher, p<0.001). Side effects were also reported more commonly by those who expected them before starting the new medication (55.4% versus 14.7%, p<0.001).

Variables entered in the multivariable logistic regression model included age, sex, and the five variables significantly associated with early non-persistence in univariate models: knowledge score, health care provider score, expectation for side effects, beliefs, and incident side effects.[Table 3] In the final model, three variables were significantly associated with non-persistence: 1) negative beliefs (i.e., overall beliefs score<11): OR 7.47, 95% CI: 5.13 to 10.86); 2) incident side effects: OR 8.00, 95% CI 5.49 to 11.68; and 3) expectations for side-effects before starting treatment: OR 0.57, 95% CI: 0.39 to 0.83.

**Table 3.** Odds ratios and 95% confidence intervals for logistic regression analysis of risk factors associated with medication non-persistence.

| Variable | Odds Ratio (unadjusted) | 95% Confidence interval |  | P value | Odds Ratio (adjusted<sup>δ</sup>) | 95% Confidence interval |  | P value |
| --- | --- | --- | --- | --- | --- | --- | --- | --- |
| Female (vs male) | 1.20 | 0.89 | 1.63 | 0.237 | 1.06 | 0.75 | 1.50 | 0.747 |
| Age 65 or older (vs less than 65 years) | 0.95 | 0.70 | 1.29 | 0.736 | 1.03 | 0.72 | 1.46 | 0.889 |
| Knowledge score < 7 (vs 7 or higher) | 2.07 | 1.53 | 2.81 | **<0.001** | 1.07 | 0.71 | 1.60 | 0.756 |
| Health Care Provider Score < 17 (vs 17 or higher) | 2.39 | 1.73 | 3.29 | **<0.001** | 1.16 | 0.76 | 1.78 | 0.487 |
| Expectation for side effects prior to starting medication (yes vs no)* | 1.61 | 1.17 | 2.21 | **0.003** | * | * | * | 0.211 |
| Beliefs score <11 (vs 11 or higher) | 9.95 | 7.22 | 13.70 | **<0.001** | 7.26 | 4.98 | 10.59 | **<0.001** |
| Side effects (yes vs no) | 8.40 | 6.03 | 11.69 | **<0.001** | * | * | * | **<0.001** |
| Interaction term (side effects and expectation for side effects before starting therapy) | - | - | - | - | - | - | - | **<0.001** |

<sup>δ</sup>Odds ratios are adjusted for all variables in the table (sex, age, knowledge score, health care provider score, expectation for side effects, beliefs score, side effects, and the two way interaction term)
\*OR suppressed due to the significant interaction between expectation for side effects and actual side effects.

No statistically significant interaction was detected between occurrence of side effects and beliefs. However, the two-way interaction between the expectation for side effects and incident side effects was significant; thus, the interaction term was included in the final model. The c-statistic for the final model containing the main effects plus the two-way interaction was 0.84.

Investigation of the significant interaction term revealed a potentially important effect modification. Among respondents with incident side effects, the risk for early non-persistence was 11.5% if they indicated an expectation for side effects before starting the medication compared to 23.6% if they did not expect side effects prior to starting the drug (adjusted OR 0.38, 95% CI 0.25 to 0.60). A prior expectation for side effects was less important for those who did not experience side effects from their new drug (i.e., 4.3% vs 2.0% for those expecting vs not expecting side effects, adjusted OR 1.59, 95% CI 0.85 to 2.98).

## Discussion

We conducted a population-based survey study of individuals receiving a new medication from one of three common pharmacologic classes: antihypertensives, cholesterol-lowering, or antihyperglycemics. Negative beliefs and incident side effects were the strongest risk factors for early non-persistence; their effects were additive but we found no evidence that they influence each other. Individuals with both side effects and negative beliefs exhibited the highest risk for non-persistence while the absence of both factors virtually eliminated the risk. In contrast to beliefs, an expectation for side effects before starting treatment appeared to protect against the negative influence of incident side effects. Specifically, those who expected side effects were half as likely to discontinue if side effects occurred. However, the majority of respondents expected no side effects before starting their new medication.

Previous studies have linked expectations for side effects with negative beliefs and the emergence of nocebo effects, while individuals expecting no side effects are considered optimistic and accepting of medication therapy.^17,18,20,21,28^ However, studies reporting associations with nocebo effects tend to be conducted in highly controlled conditions, often with healthy volunteers responding to hypothetical situations.^28^ Our population was drawn from real world patients initiating a medication for a chronic condition. Although we found a higher incidence of side effects among those who expected them, the results of our study provide a very different perspective about the relevance of expectations for side effects. Expecting no side effects before starting a new medication appeared to be an inconspicuous risk factor for non-persistence. Perhaps expectations for side effects represented a level of preparedness or commitment to a new medication that anticipated possible tolerability challenges. Although further research is required to understand the nature of this association and the potential role health care providers might play, it appears reasonable to for practitioners to ensure patients understand that side effects are a real possibility, which may be ignored if patient education is overly optimistic. At minimum, frank conversations about accepting negative medication effects may lead to a better understanding of a patient’s level of commitment to a new treatment.

Three quarters of our respondents did not expect side effects from their new drug. It is not immediately clear how to reconcile the low numbers of people expecting side effects compared with widespread public awareness of drug adverse effects^29^ and the attention given to patient education from health care providers.^22^ Accepting that public concern about prescription drug safety is extensive, we do not believe our findings reflect a general lack of awareness or knowledge about drug side effects. Rather, it appears that many patients may be unprepared for the emergence of side effects from new medications despite information they *might* occur. It is not clear whether expectations play a causal role in protecting against non-persistence or if they indicate other factors as yet unidentified. Regardless, expectations for side effects may be relatively simple to evaluate and could have far more prognostic importance that previously thought.

### Limitations

We conducted a rigorous analysis of patient-reported risk factors for non-persistence. However, several limitations must be noted. First, our data was derived from voluntary responders to a population-level study invitation. Thus, we cannot be certain the aggregate experiences collected from our study sample reflect population averages. For example, the low frequency of negative beliefs observed in our sample may have been influenced by a healthy responder bias.^30^ Despite this limitation, we found clear associations between negative beliefs and side effects with non-persistence to medications that we believe exist in the general population. Second, our questionnaire was developed for maximum breadth of potential adherence risk factors. To minimize responder fatigue, we collected important factors such as negative beliefs using abbreviated measures in many cases. Our approach produced two important disadvantages, it reduced the potential granularity of our analysis relating to the major sub-scales of the beliefs paradigm (i.e., necessity, concern, overuse, and harm);^31^ it also potentially reduced the validity of the measure given the reliance on a very limited number of questions. However, we had high confidence in the clarity and face validity of our beliefs questions. Also, the association between our questions about side effects, beliefs, and expectations were very strong and allowed for a clear assessment of impact and interaction with a wide range of other factors. Third, the cross-sectional design of our study made it impossible to confirm temporal relationships between beliefs, starting the new drug, and emergence of side effects. Thus, we can only speculate about the true nature of the associations reported in this study. Finally, all our data were self-reported. Thus, we cannot verify the accuracy and honesty of respondents to vital questions such as “*Are you still taking the medicine prescribed to you*” (i.e., our primary endpoint). However, we have confidence in the clarity and importance of discontinuing medication as a definitive and relevant outcome.

## Conclusion

Prior to starting a new medication, an individual’s expectations for side effects may be a previously unrecognized but important marker of their probability to persist with treatment. A high percentage of new medication users appear unprepared for the possibility of side effects from their new medication making them less resilient if side effects occur.

### Transparency statement

David Blackburn (the lead author) affirms that the manuscript is an honest, accurate, and transparent account of the study being reported; no important aspects of the study have been omitted; and any discrepancies from the study as originally planned have been explained.

### Role of the funding source

This study was funded from a peer reviewed research grant from the Canadian Institutes of Health Research (CIHR grant number 130343). CIHR had no role in the study design, collection, analysis, interpretation of data, writing of the report, or in the decision to submit the article for publication. In addition, all authors had full access to all of the data (including statistical reports and tables) in the study and can take responsibility for the integrity of the data and the accuracy of the data analysis.

### Dissemination to participants and related patient and public communities

We did not collect identifiable information on respondents; thus, results cannot be shared directly.

## Data Availability

The data that support the findings of this study are available from the corresponding author upon reasonable request.

## Notes

### Competing Interest Statement

The authors have declared no competing interest.

### Author Declarations

Our study was approved by the University of Saskatchewan Committee for Ethics in Human Research.

